# Population pharmacokinetics and target attainment of pretomanid in rifampicin-resistant Tuberculosis patients

**DOI:** 10.64898/2025.12.04.25341619

**Authors:** Bern-Thomas Nyang’wa, Ilaria Motta, Ronelle Moodliar, Varvara Solodovnikova, Shakira Rajaram, Mohammed Rasool, Catherine Berry, Zhonghui Huang, Geraint Davies, David Moore, Frank Kloprogge

**Author notes:** Corresponding authors Bern-Thomas Nyang’wa; Frank Kloprogge.

## Abstract

Pretomanid is a key component of the bedaquiline, pretomanid, linezolid with or without moxifloxacin (BPaL/M)regimen recommended for treatment of rifampicin-resistant tuberculosis (RR-TB). To support dose optimization and efficacy interpretation, we developed a pretomanid population pharmacokinetic (PK) model and evaluated exposure and probability of target attainment (PTA). Ninety-four RR-TB patients received daily oral pretomanid at 200 mg, and plasma samples were collected at multiple time points. Pretomanid concentrations were quantified using high-performance liquid chromatography-tandem mass spectrometry and PK modeling was performed using nlmixr2 in R. A one-compartment model with first-order absorption and elimination, and fat free mass allometric scaling best described the data. Typical clearance was 3.10 L/h, median AUC₀₋₂₄ was 63,733 μg·h/L, and median trough concentration was 1,965 μg/L. Pretomanid MICs for *Mycobacterium tuberculosis* in the TB-PRACTECAL trial were consistently below the provisional critical concentration, with a median of 0.125 mg/L. PK-Pharmacodynamic (PD) simulations indicated that nearly all participants achieved drug exposures exceeding %fT>MIC, supporting the regimen’s efficacy across the study population. We developed a pretomanid population PK model and facilitated exploring robust PK-PD targets for PTA that remain valuable to support dose optimisation. There is an urgent need for further research to identify the optimal clinically relevant PK-PD index for pretomanid, especially within the context of combination therapy.

## Introduction

Tuberculosis (TB) is the leading cause of death from a single infectious agent, claiming approximately 1.2 million lives globally each year. The emergence of drug-resistant strains poses a significant challenge to TB control efforts(1). The World Health Organization recommends a six-month, fully oral regimens comprising bedaquiline, pretomanid and linezolid, with or without moxifloxacin (BPaL/M) as the preferred treatment approach for rifampicin-resistant tuberculosis (RR-TB)(2). These regimens and together with bedaquiline, pretomanid, linezolid, and clofazimine (BPaLC), were investigated for the treatment of RR-TB in the TB-PRACTEAL randomized controlled clinical trial(3, 4).

Pretomanid is a nitroimidazooxazine antimycobacterial approved for the treatment of TB as part of a BPaL regimen. It is extensively metabolized through reductive and oxidative metabolism but no major pathway has been identified thus far. Around 20% is metabolised through cytochrome P450-3A and 1% appears in urine as unchanged pretomanid. At an oral dose of 200 mg, steady state pharmacokinetic (PK) parameters are as follows: Cmax 1.7 mg/L, Tmax of 4.5 hours, T1/2 16 hours. A high-fat, high-calorie meal increases Cmax by 76% and AUC by 88% when compared to the fasting state(5–8).

Pretomanid inhibits mycolic acid biosynthesis, thus disrupting cell wall production in actively replicating *Mycobacterium tuberculosis*. It also kills non-replicating bacteria in anaerobic environments by generating reactive nitrogen species including nitric oxide(9). Maximum efficacy is achieved with a daily dose of 200mg, increased toxicity but not increased efficacy is experienced with higher doses in early bactericidal studies(10); doses of 50mg, 100mg and 150mg have captured the incremental efficacy and toxicity better(11). At a dose interval of less than 48 hours, pretomanid efficacy PK-Pharmacodynamic (PD) can be defined by area under the free drug concentration curve (fAUC/MIC) or cumulative percentage of the dosing interval that the free drug concentration exceeds the MIC (fT>MIC) parameters(12). However, pretomanid protein binding *in vivo* has not yet been determined, approaches have included using total drug concentration, 85% binding(7, 12, 13).

This study aimed to investigate pretomanid PK characteristics amongst rifampicin resistant tuberculosis patients in the TB-PRACTECAL trial receiving combination therapy and to assess previously reported PK-PD endpoints against prospectively collected *Mycobacterium tuberculosis* susceptibility data.

## Results

36% of the 94 study participants were female and the median age was 36 years (range: 19 - 71 years) (Table 1). The total number of timed pretomanid plasma samples included in the population PK analysis was 952, 86 samples were collected before the first dose and 866 samples after the first dose. A total of 234 samples were below the limitation of quantification (BLQ), of which 151 samples were collected after treatment completion, rendering 9.5% of the pretomanid concentration BLQ during treatment. Observed pretomanid plasma concentrations ranged between 19.1 and 11,566 ng/ml, the median trough concentration was 1,789 ng/ml, with an interquartile range of 1,126 – 2,689 ng/ml.

**Table 1:** Baseline characteristics of the study participants.

|  |  |
| --- | --- |
| Sample size (n) | 94 |
| Age (y) | 36 (19-71) |
| Baseline weight (kg) | 56.8 (39.2-144.4) |
| Body mass index (kg/m<sup>2</sup>) | 19.7 (14.3-47.1) |
| Fat free mass (kg) | 45.5 (28.6-75.5) |
| BUN (mmol/L) | 3.6 (1.7-8.5) |
| ALT (IU/L) | 19.5 (4-113) |
| AST (IU/L) | 22 (4-82) |
| Total protein (g/L) | 77 (61-118) |
| Estimated creatinine clearance (mL/min) | 105.4 (43.4-243.8) |
| Female, n (%) | 34 (36.2) |
| Caucasian race, n (%) | 40 (42.6) |
| Black race, n (%) | 52 (55.3) |
| Asian race, n (%) | 1 (1.1) |
| Other race, n (%) | 1 (1.1) |
| HIV positive, n (%) | 39 (41.5) |
| BPaLM, n (%) | 38 (40.4) |
| BPaLC, n (%) | 30 (31.9) |
| BPaL, n (%) | 26 (27.7) |

Median (min-max) if not stated otherwise. BUN = Blood urea nitrogen, ALT= alanine transaminase, AST= aspartate aminotransferase.

A one-compartment first order absorption and elimination model best described the observed pretomanid PK time series data. Random effects on clearance and central volume of distribution explained inter-individual variability and a combined residual error model was used to characterise the unexplained variability (Table 1 and appendix 1).

Allometric body size scaling on clearance and volume of distribution, using fat free mass, was embedded *a priori*. Step-wise covariate modelling rendered female gender, black race, and ‘BPaL regimen’ on volume of distribution in forward first step analysis (p < 0.05, per degree of freedom). However, none of these were significant in the backward elimination step (p < 0.001, per degree of freedom) (Appendix 2).

Goodness-of-fit plots for the pretomanid population PK model showed no significant bias from the unity line, indicating that the model predicted individual and population values closely matched the observed PK data (Figure 1). Model validation using a visual predictive check (VPC) and individual model fit, confirmed the predictive accuracy of the pretomanid model (Figure 2 and Appendix 3). Pretomanid population PK model and secondary parameters are shown in Table 2 and Table 3.

**Figure 1:**
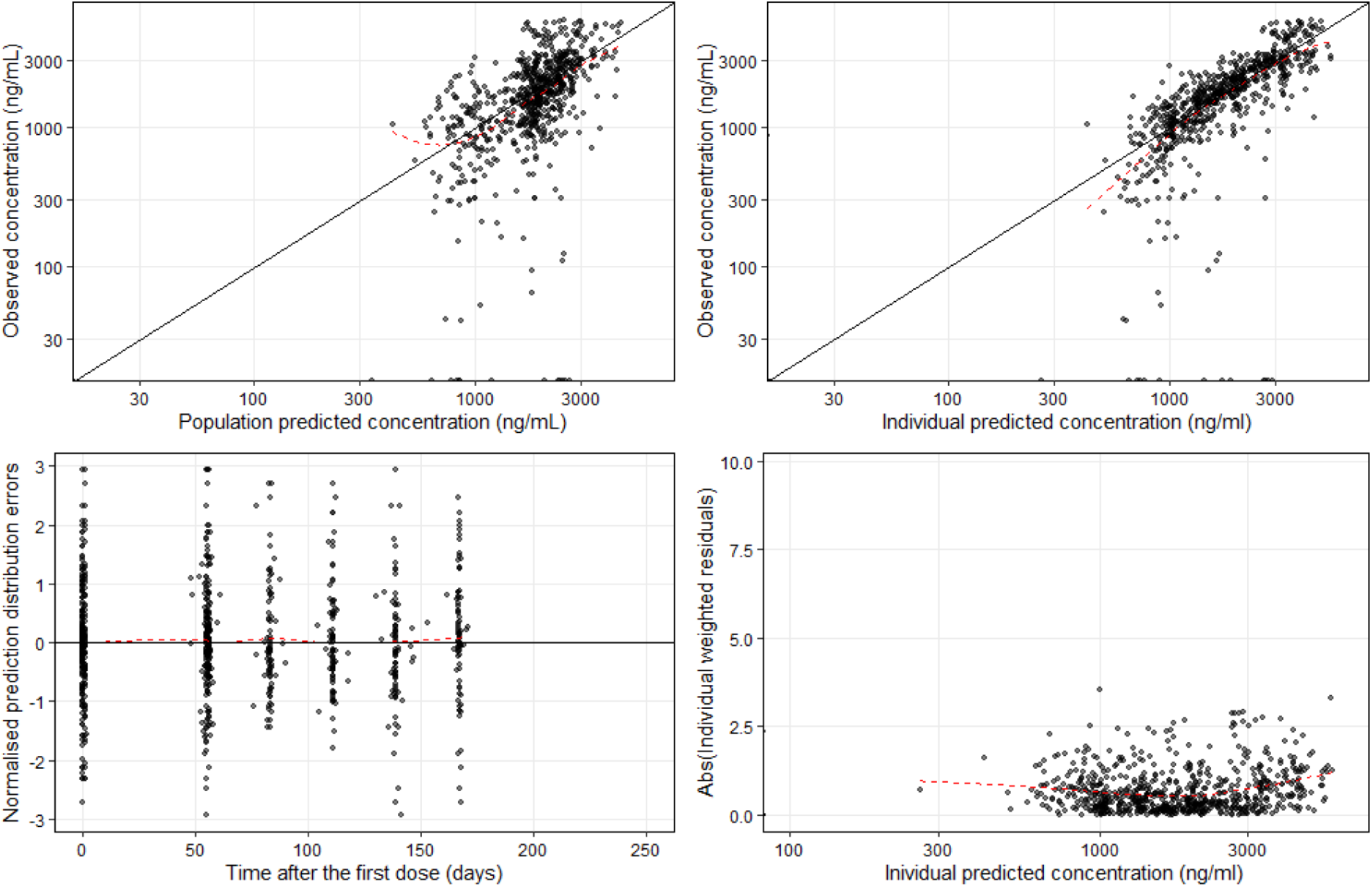
Pretomanid population pharmacokinetic model goodness of fit plots. Dots represent observations, solid black lines represent identity lines and the red dashed lines represent locally estimated scatterplot smoothing.

**Figure 2:**
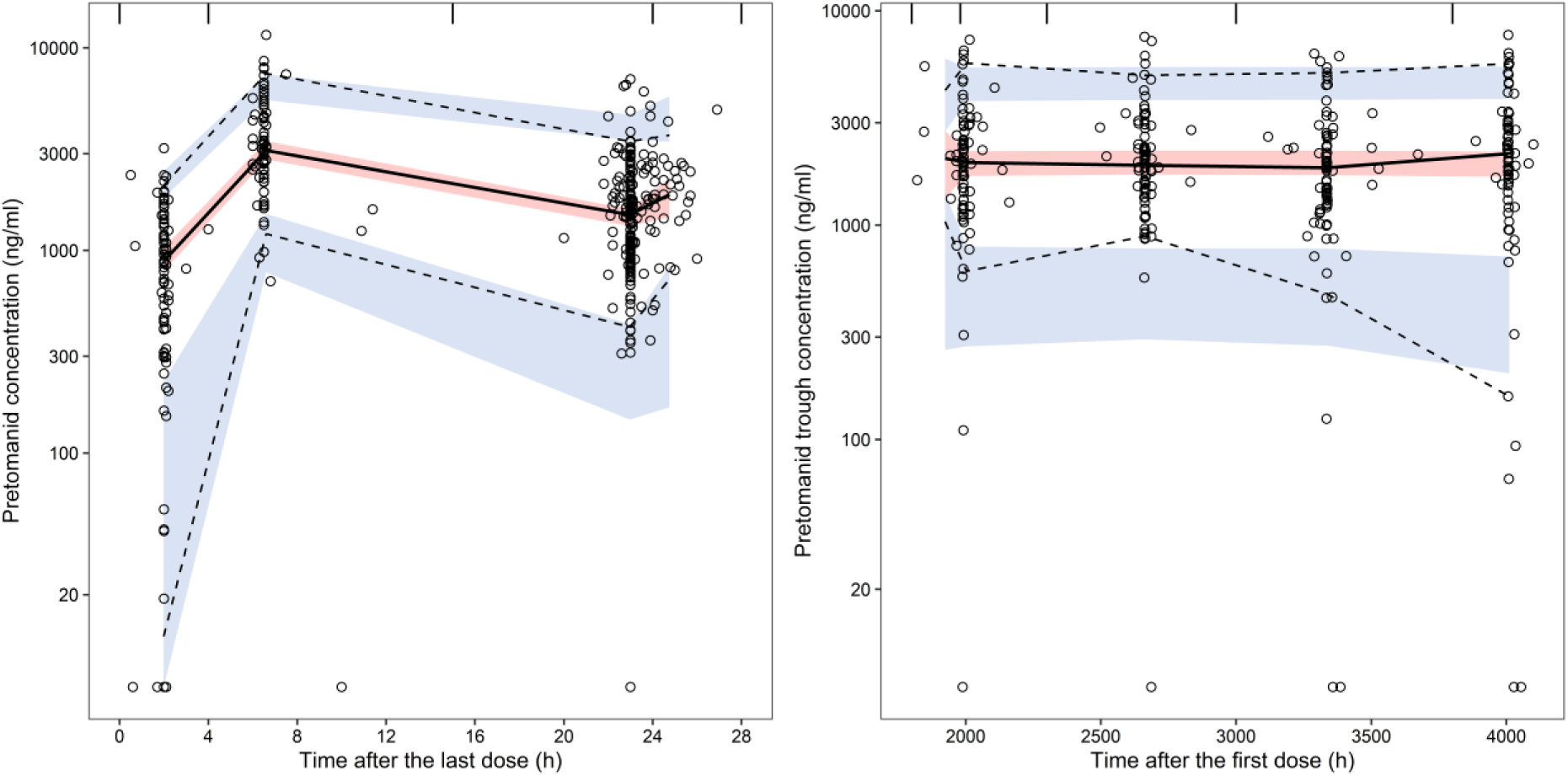
Visual predictive check, presented as time after last dose of the day 1 and week 8 visit (left panel) and time after first dose for the subsequent visits (right panel). Open circles represent the observed data, with dashed and solid lines presenting the 95% percentiles and median of the observed data. The blue and red shaded areas represent the 90% confidence intervals of model predicted 95% percentiles and median of the simulated data.

**Table 2:**
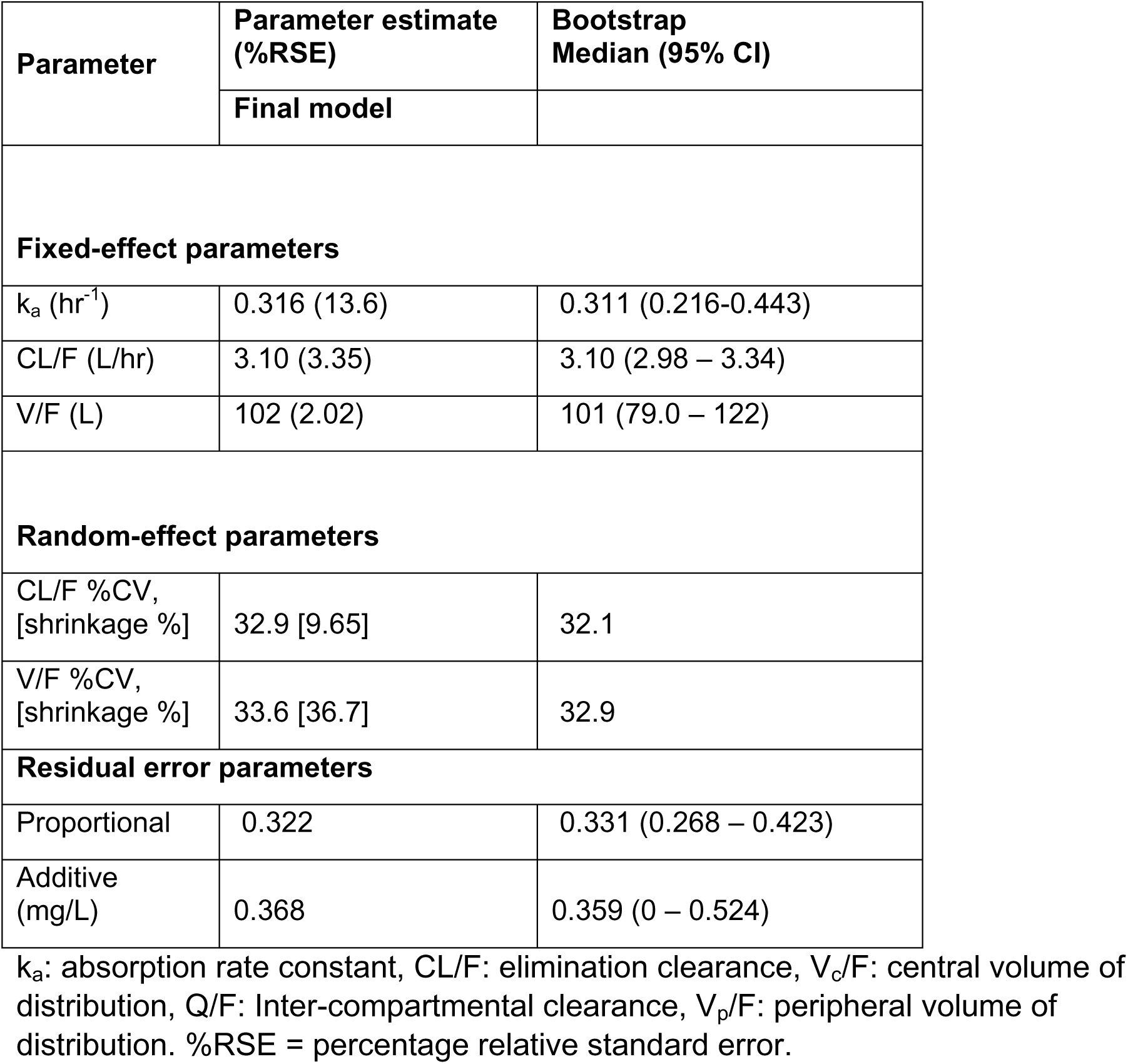
Estimated pretomanid population pharmacokinetic parameters.

| Parameter | Parameter estimate<br>(%RSE) | Bootstrap<br>Median (95% CI) |
| --- | --- | --- |
|  | Final model |  |
| **Fixed-effect parameters** |  |  |
| $k_a$ ($\text{hr}^{-1}$) | 0.316 (13.6) | 0.311 (0.216-0.443) |
| CL/F (L/hr) | 3.10 (3.35) | 3.10 (2.98 – 3.34) |
| V/F (L) | 102 (2.02) | 101 (79.0 – 122) |
| **Random-effect parameters** |  |  |
| CL/F %CV,<br>[shrinkage %] | 32.9 [9.65] | 32.1 |
| V/F %CV,<br>[shrinkage %] | 33.6 [36.7] | 32.9 |
| **Residual error parameters** |  |  |
| Proportional | 0.322 | 0.331 (0.268 – 0.423) |
| Additive<br>(mg/L) | 0.368 | 0.359 (0 – 0.524) |

$k_a$ : absorption rate constant, CL/F: elimination clearance, $V_c/F$ : central volume of distribution, $Q/F$ : Inter-compartmental clearance, $V_p/F$ : peripheral volume of distribution. %RSE = percentage relative standard error.

**Table 3:**
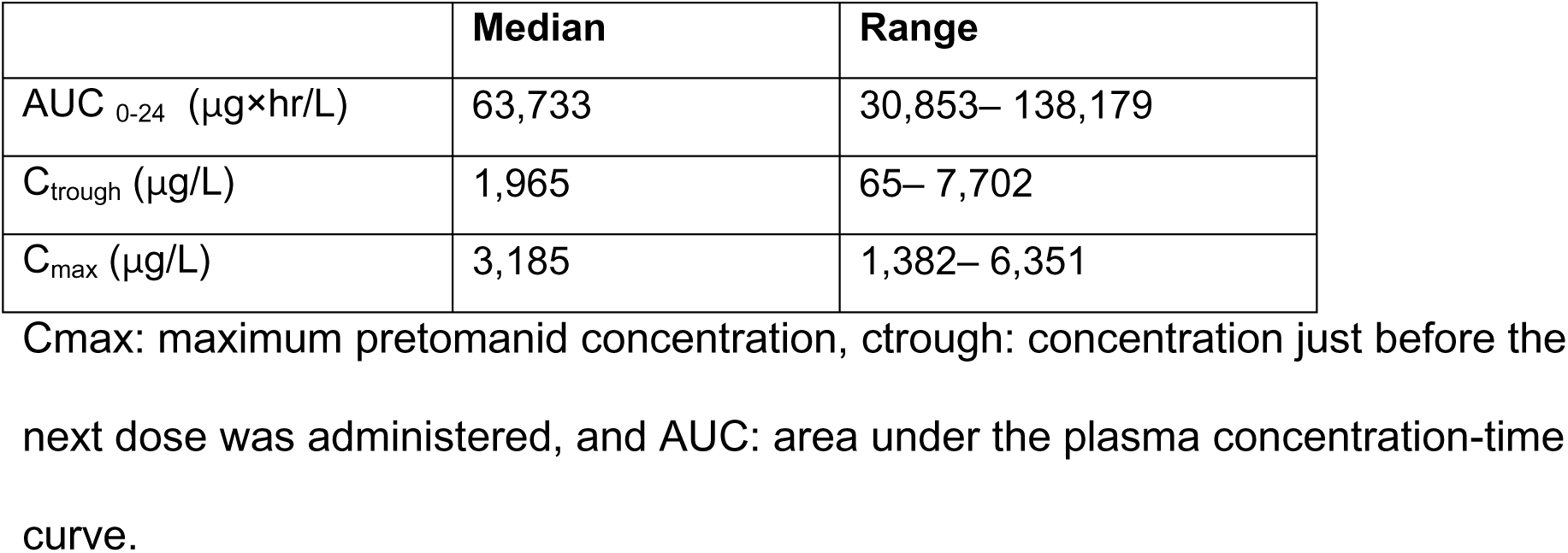
Secondary pharmacokinetic parameters derived with the pretomanid model.

The distribution of *Mycobacterium tuberculosis* MICs for pretomanid tested using MGIT from 478 TB-PRACTECAL study participants was disaggregated by country of enrolment (Figure 3). The median MIC was 0.125mg/L and the interquartile range from 0.125 to 0.25mg/L. 100% of the isolates were below the provisionally set critical concentration of 1mg/L(14) or 2mg/L(15). Only two isolates, both from South African sites had a baseline MIC of 1.0mg/L.

**Figure 3:**
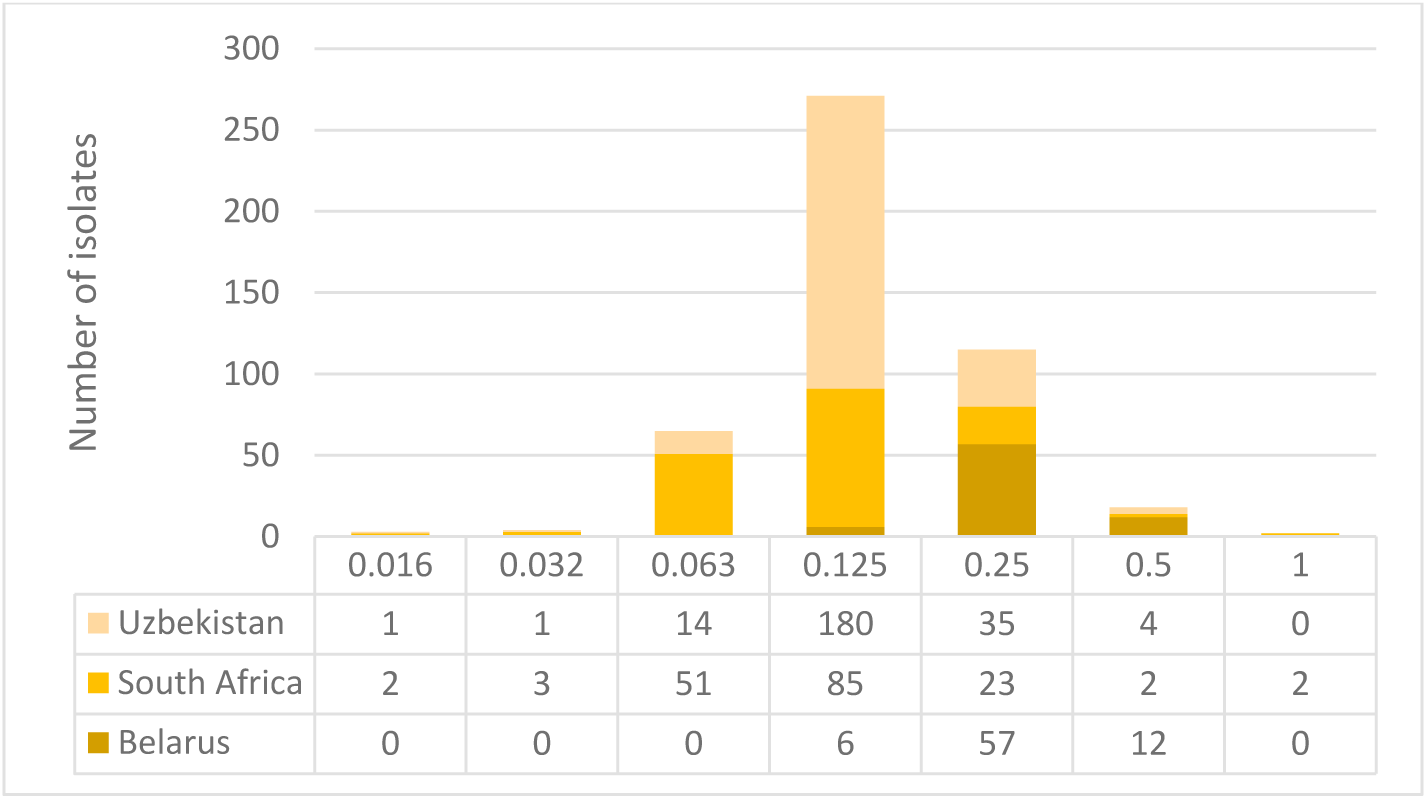
Distribution of *Mtb* baseline isolates across various pretomanid MICs (mg/L) in the TB-PRACTECAL trial.

Assuming 85% plasma protein binding, a 200 mg dose resulted in 80% or higher PTA fAUC0-24/MIC of 167 at an MIC of 0.032mg/L and below (Figure 4). PTA stochastic simulations (n=2,000) using fT>MIC showed that both target of 77% and 48%, associated with 1 and 1.59 log10 kill, were met at 200 mg dosing at MICs of 0.125 and 0.250 mg/L (Figure 4). Only at the lowest MIC of 0.032mg/L would a 200 mg dose attain both fAUC0-24/MIC and fT>MIC targets.

**Figure 4:**
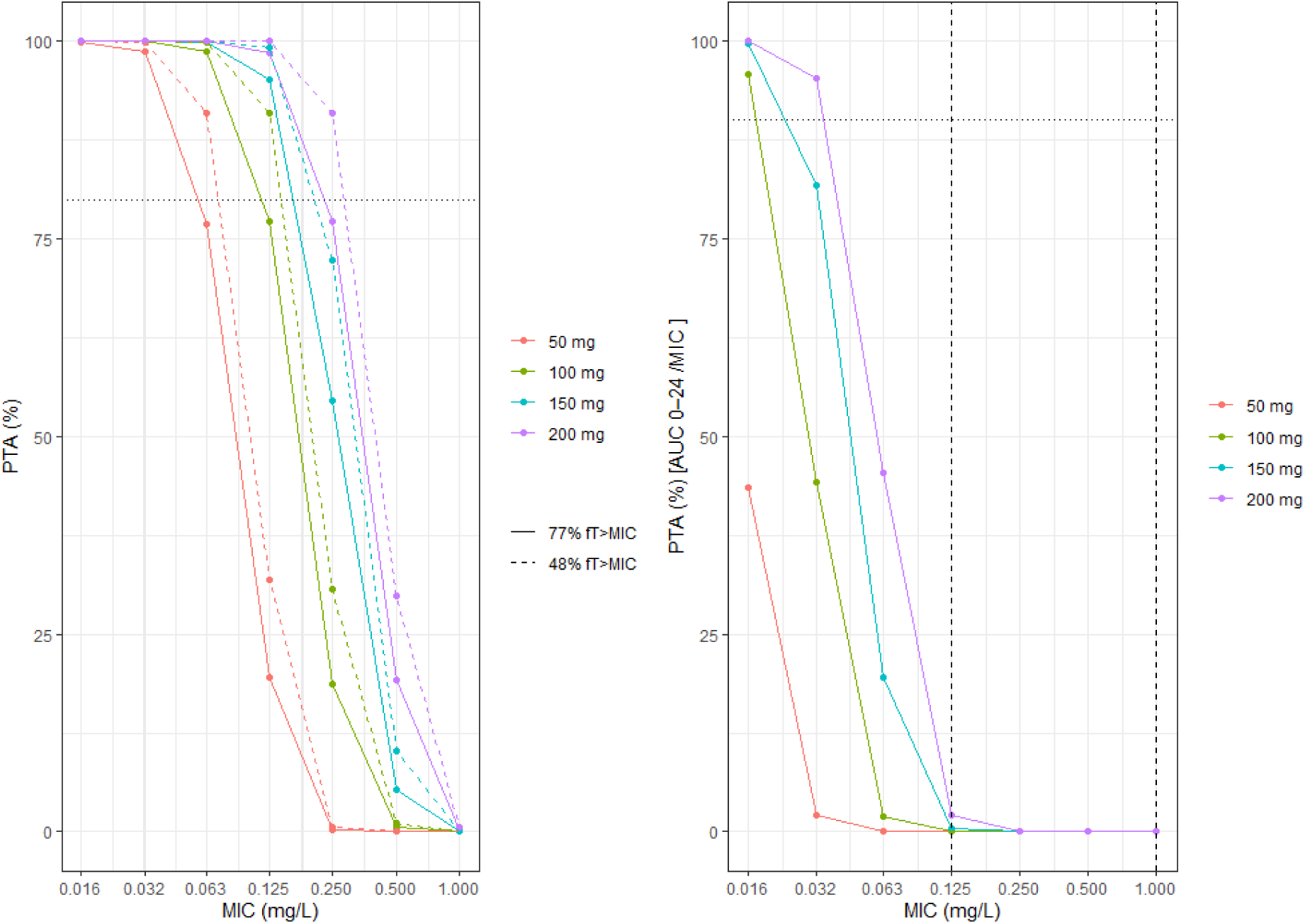
PTA plot for day 14 fT%>MIC (left) and fAUC_0-24_/MIC (right). Both assumed 85% plasma protein binding and a PK-PD target of 167 was selected for fAUC_0-24_/MIC.

Given the pretomanid median MIC at baseline in the study population was 0.125 mg/L, virtually all patients in the TB-PRACTECAL trial had drug exposures above %fT>MIC targets, for the fAUC/MIC target this dropped to near 0%.

## Discussion

A population pharmacokinetic model for pretomanid was developed using data from the largest single-study cohort to date, which included participants from both South Africa and Belarus. In contrast, previous studies, limited to a single country, only investigated dosing durations of up to six months. These results also confirm the adequate pretomanid exposure in rifampicin resistant TB regimens of BPaLM and BPaL which are the currently recommended regimens by WHO(2).

A one-compartment first order absorption and elimination model with allometric scaling of fat-free mass on both clearance and volume of distribution best described pretomanid PK characteristics. The clearance in our study was estimated at 3.10 L/hr, this is similar to clearances which have been previously published(7, 16–19).

The only identified covariate in our study was fat-free mass (Table 2), concurring with the allometry body size scaling that has been previously reported in other studies using weight(7, 16) and FFM(18).

Food administration, particularly high calorie, high fat meal, is postulated to significantly increase pretomanid exposure(8, 18). However, other studies have found the fed state to influence absorption but not bioavailability and consequently exposure(19). Our study participants were encouraged to eat before taking medicine. There was no standardised meal, observation, or recording of the type of meal consumed, and therefore current exposures are expected to be representative of real-world scenarios.

Co-administration of potent CYP450 inducers such as efavirenz, lopinavir/ritonavir and rifamycins reduce pretomanid exposure(6, 18). However, these drug classes were contraindicated in the TB-PRACTECAL trial. All the 39 participants living with HIV (42% of total study participants) were on integrase inhibitors and nucleotide reverse transcriptase inhibitor antiretroviral regimens; since HIV was not a significant covariate no further exploration of individual drugs’ effect on the PK model was done. Both moxifloxacin and clofazimine are metabolised in the liver(20, 21). However, including BPaLM and BPaLC as covariates on clearance did not improve the model fit significantly, suggesting none or limited impact of the accompanying anti-TB drugs in the regimen on pretomanid exposure.

In our study 72% of the participants had an MIC of equal to or lower than 0.125 mg/L. Whilst the MIC in this study was measured using MGIT, pretomanid MIC on solid phase media Middlebrook 7H11 has been reported within a similar range at 0.03125 to 0.25 mg/L(22). At the standard 200mg daily dose that was used in the TB-PRACTECAL study, assuming 85% protein binding, all isolates within with MIC of 0.250 mg/L or lower, making up 95% of the participants in TB-PRACTECAL, would have achieved the maximal efficacy %fT>MIC targets for a 1-log10 kill (Figure 4). When using fAUC/MIC as PK-PD target at 167 and the trial’s dose of 200mg daily, adequate exposure would only be achieved for strains with an MIC of 0.032 mg/L or below (Figure 4). Further studies to establish the best pretomanid clinical PK-PD index, preferably using MGIT MIC and when given as part of combination treatment, are therefore urgently needed.

Limitations of the study included the lack of closely observing food intake. Our study may therefore under- or over-estimate pretomanid drug exposure, although this has previously been shown not to be clinically significant(8). The trial excluded patients with moderate liver and renal function abnormality, which makes it less representative of the full spectrum of RR-TB patients.

Pretomanid when given at 200mg daily in combination with bedaquiline, linezolid with/without moxifloxacin or clofazimine results in adequate exposure. This is the case also in HIV coinfected patients taking antiretroviral treatment consisting of integrase inhibitors and nucleoside reverse transcriptase inhibitors. Assumed protein binding and type of PK-PD index alter the interpretation of probability of target attainment significantly. Further studies to establish pretomanid protein binding in humans to inform an optimal PK-PD PTA index and target are recommended.

## Materials and Methods

Pretomanid concentration time series data was available from a nested pharmacokinetic-pharmacodynamic study in TB-PRACTECAL, a randomized controlled trial with rifampicin resistant tuberculosis patients(3, 4, 23). All three investigational study arms contained pretomanid as part of the BPaL backbone which was administered alone or with moxifloxacin or clofazimine. Bedaquiline was administered daily for 2 weeks at 400mg, then at 200mg three times a week for 22 weeks, linezolid was administered daily at 600mg for 16 weeks, then at 300mg daily for the remaining 8 weeks, whilst pretomanid was administered at 200mg daily for 24 weeks. Clofazimine in the BPaLC arm was administered at 100mg daily for 24 weeks and moxifloxacin was administered at 400mg daily for 24 weeks in BPaLM arm.

Blood samples for pretomanid plasma concentration quantification were collected on Day 1 (0, 2 and 23 hours), Weeks 8 (predose, 6.5 and 23 hours), 12, 16, 20, 24, 32 and 72 post randomisation visits. Pretomanid concentrations were quantified in a GCP laboratory using a high-performance liquid chromatography-tandem mass spectrometry. The lower limit of quantification for pretomanid was 7ng/mL.

All data manipulations and analyses were done in R v4.1.2 using nlmixr2, an open-source packages for nonlinear mixed-effect modelling. A list of all R-pakages is listed in appendix 5. Pretomanid concentration time series data were fitted using a first-order conditional estimation with interaction (FOCE-I) algorithm, whilst Intir-individual variability (IIV) at parameter level described variability between patients and residual variability (RV) at observation level described random variability.

Most pretomanid population PK models in the public domain use one-compartment to describe their data(7, 16–19). Consequently, one and two-compartment linear models were evaluated using combined, proportional, additive and log-transformed residual error models. Estimated and fixed first order and transit absorption models were tested to describe pretomanid absorption and random effects on elimination clearance and distribution volume were evaluated.

A matrix comprising covariates and individual eta estimates for clearance and volume of distribution from the base model was used to assess both correlation and collinearity among variables. Allometric body size scaling was applied to clearance and volume parameters, with fixed exponents of 0.75 for clearance and 1 for volume. Covariate selection was performed using a stepwise approach: forward inclusion was based on a significance threshold of p < 0.05 (ΔOFV > 3.84 per degree of freedom), followed by backward elimination using a stricter criterion of p < 0.001 (ΔOFV > 10.83 per degree of freedom) to retain only those covariates that significantly enhanced model performance.

Goodness-of-fit plots were used to assess how well the model predicted individual and population values closely matched the observed PK data. Model validation was also performed using visual predictive check (VPC) plots. The shrinkage, relative standard error, and variability value including omega and sigma value were also used to assess the precision and robustness of the model.

Minimum inhibitory concentrations were determined using a routine testing concentration set (1, 0.5, 0.25, 0.125, 0.063, 0.032 mg/L) in MGIT. Testing was performed using a higher (8, 4, 2 mg/L) or lower (0.016 mg/L) testing concentration set if required. The results from all participants from the TB-PRACTECAL trial were summarised by country of enrolment and the median and interquartile range reported.

PTA of pretomanid PK-PD thresholds at doses of 50mg, 100mg, 150mg and the TB-PRACTECAL dose of 200mg daily were evaluated using 2,000 Monte Carlo simulations and patient level characteristics from the observed participants in this study, accounted for protein binding at 85%-95%(7, 12, 13). The fAUC/MIC target was set at 167, which is associated with >2 log10 reduction in CFU counts(18). The fT%>MIC targets required for bacteriostatic, 1-log10 kill and 1.59- log10 kill (80% maximum effect), at 22%, 48%, and 77%, were also investigated(7).

## Data Availability

All data produced in the present study are available upon reasonable request to the authors

## Acknowledgements

The study was funded by Médecins sans Frontières. FK conducted the research as part of a Sir Henry Dale Fellowship jointly funded by the Wellcome Trust and the Royal Society (Grant Number 220587/Z/20/Z).

## Contributors

Conception and design: B-TN, FK, GD and DAM. Data acquisition: IM, RM, VS, SR. PK modelling: BT-N and ZH. First draft of manuscript: BT-N. Revising and approval of manuscript: all

## Ethics statement

The study was approved by the MSF Ethics Review Board (reference no. 1541) and the LSHTM Ethics Committee (reference no. 16249). The Belarus RSPCPT ethics committee and the regulator-Centre of Excellence for the Minsk site. PharmaEthics for the Don Mckenzie and Dorris Goodwin hospitals sites, University of Witwatersrand Human Research ethics committee for the Helen Joseph and King DiniZulu Hospitals sites and the South Africa Health Products Regulatory Authority

## Data access statement

Deidentified data will be available to researchers upon a written request to the Medical Director, Médecins sans Frontières, Operational Centre Amsterdam, the Netherlands.

## Appendix 1: R code for the pretomanid model

\# Pretomanid population pharmacokinetic model code (nlmixr2)

ini({

lka <- -1.15259732127294

label("Absorption rate")

lcl <- 1.13008124732802

label("Clearance")

lvc <- 4.62170931536226

label("Central volume of distribution")

prop.err <- c(0, 0.321731821549103)

add.err <- c(0, 367.831309683669)

covffmPow1 <- fix(0.75)

covffmPow2 <- fix(1)

eta.cl ~ 0.102674627530168

eta.vc ~ 0.10705836808838

})

model({

ka <- exp(lka)

cl <- exp(lcl + eta.cl + logFFM * covffmPow1)

vc <- exp(lvc + eta.vc + logFFM * covffmPow2)

linCmt() ~ prop(prop.err) + add(add.err)

})

## Appendix 2: Continuous covariate and parameter correlation matrix

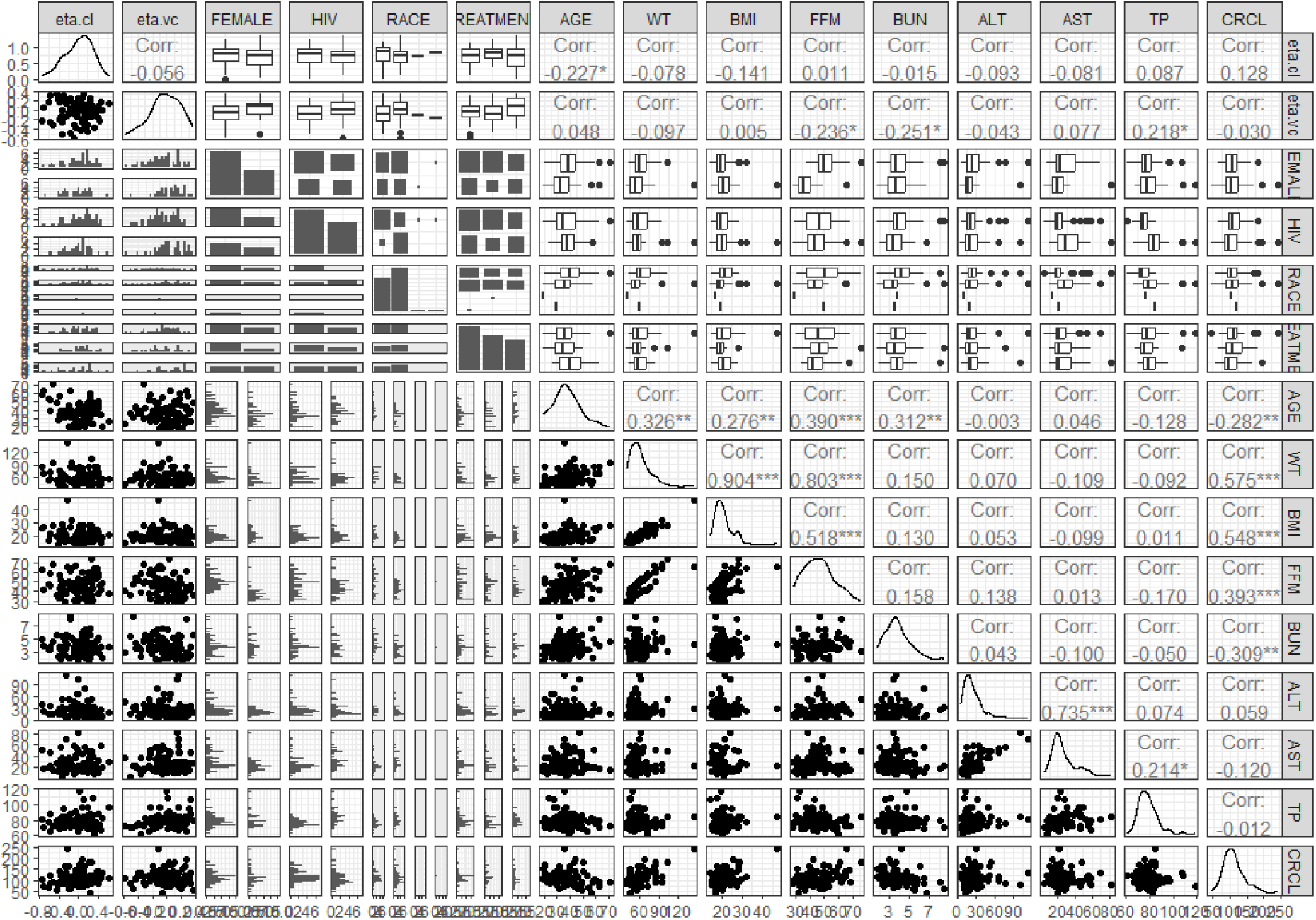

Covariate vs parameter matrix on base model. WT: weight, BMI: body mass index, FFM: fat free mass, BUN: blood urea nitrogen, ALT: alanine transaminase, and AST: aspartate aminotransferase, TP: total protein, CRCL: estimated creatinine clearance, random.g: BPaLM, BPaLC or BPaL arm.

## Appendix 3: Individual model fit plots

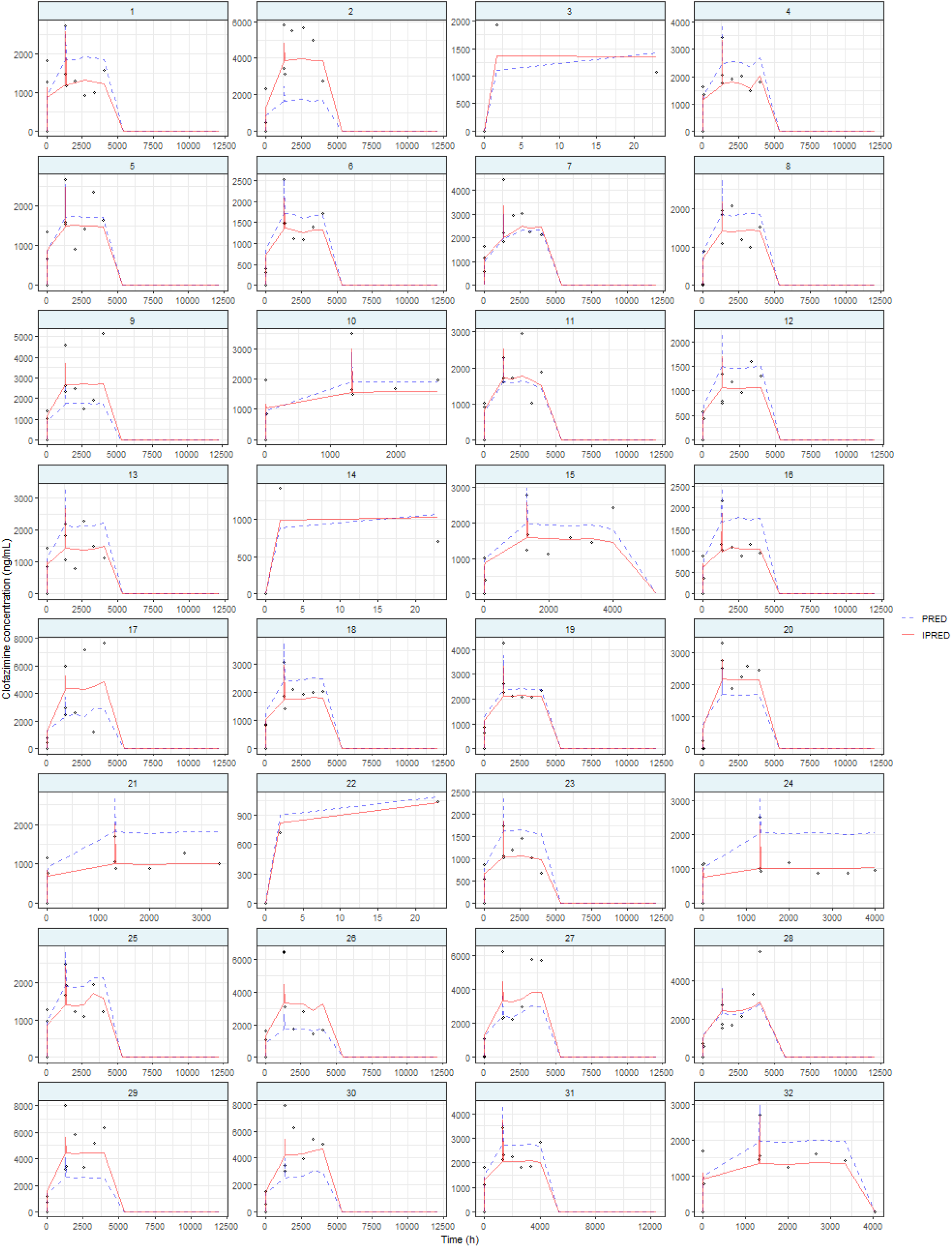
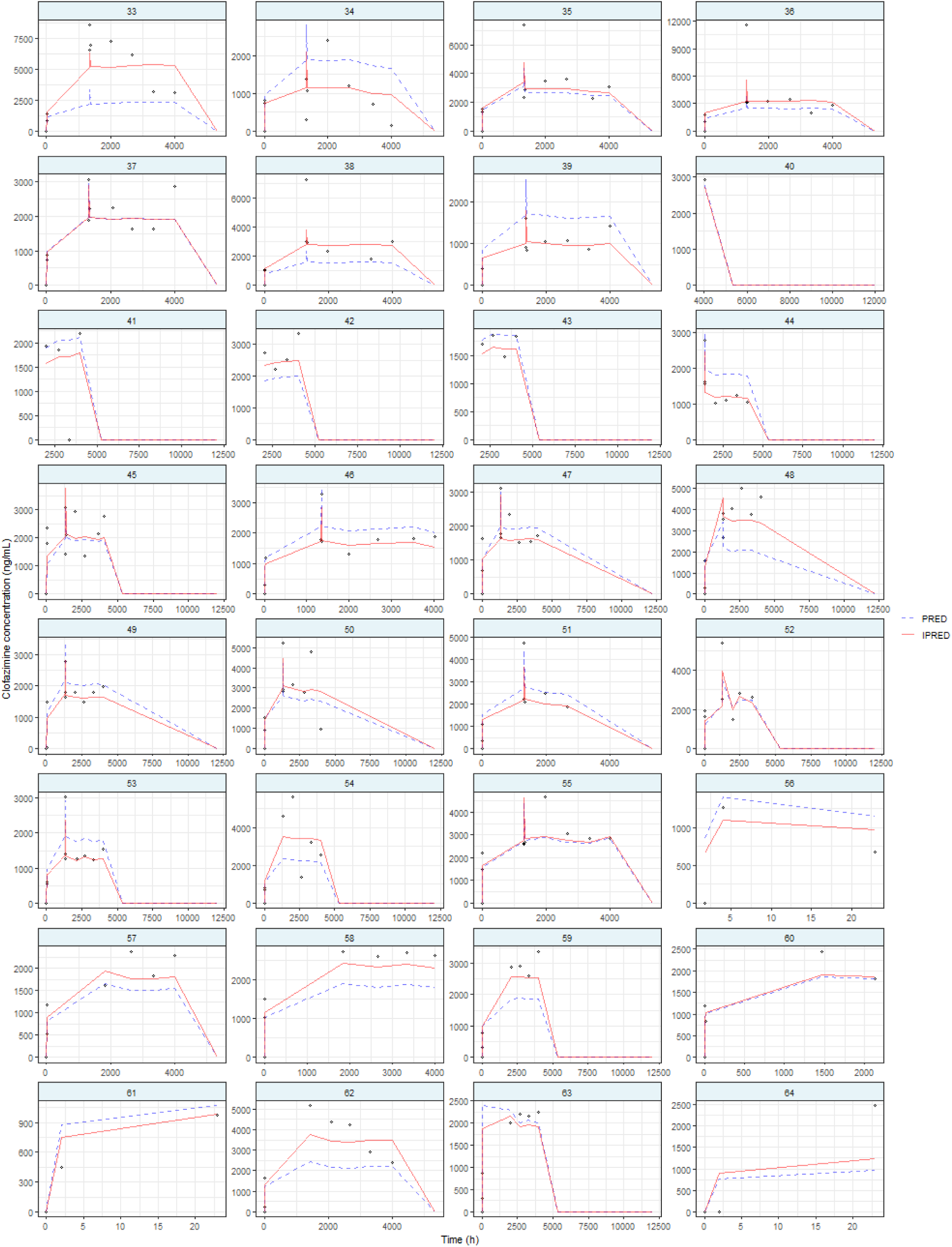
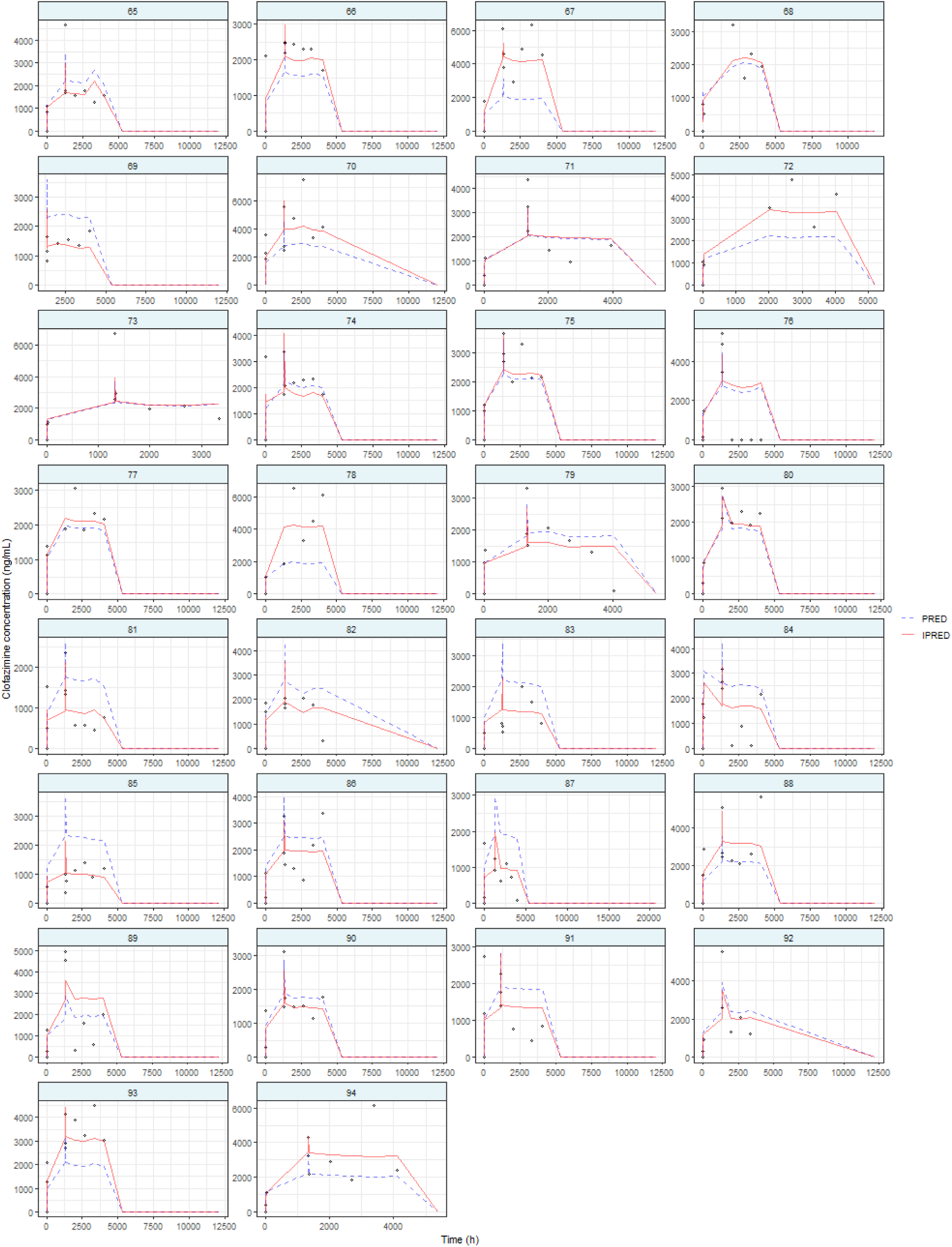

Individual linezolid plasma concentration - time profiles. Each panel represent a patient, with open circles representing observed plasma concentrations, the blue dashed line the population predictions by the developed model and the red solid lines the individual population predictions.

## Appendix 4: PTA plots for a 200 mg pretomanid dose at escalating MIC’s using varying plasma protein binding scenarios

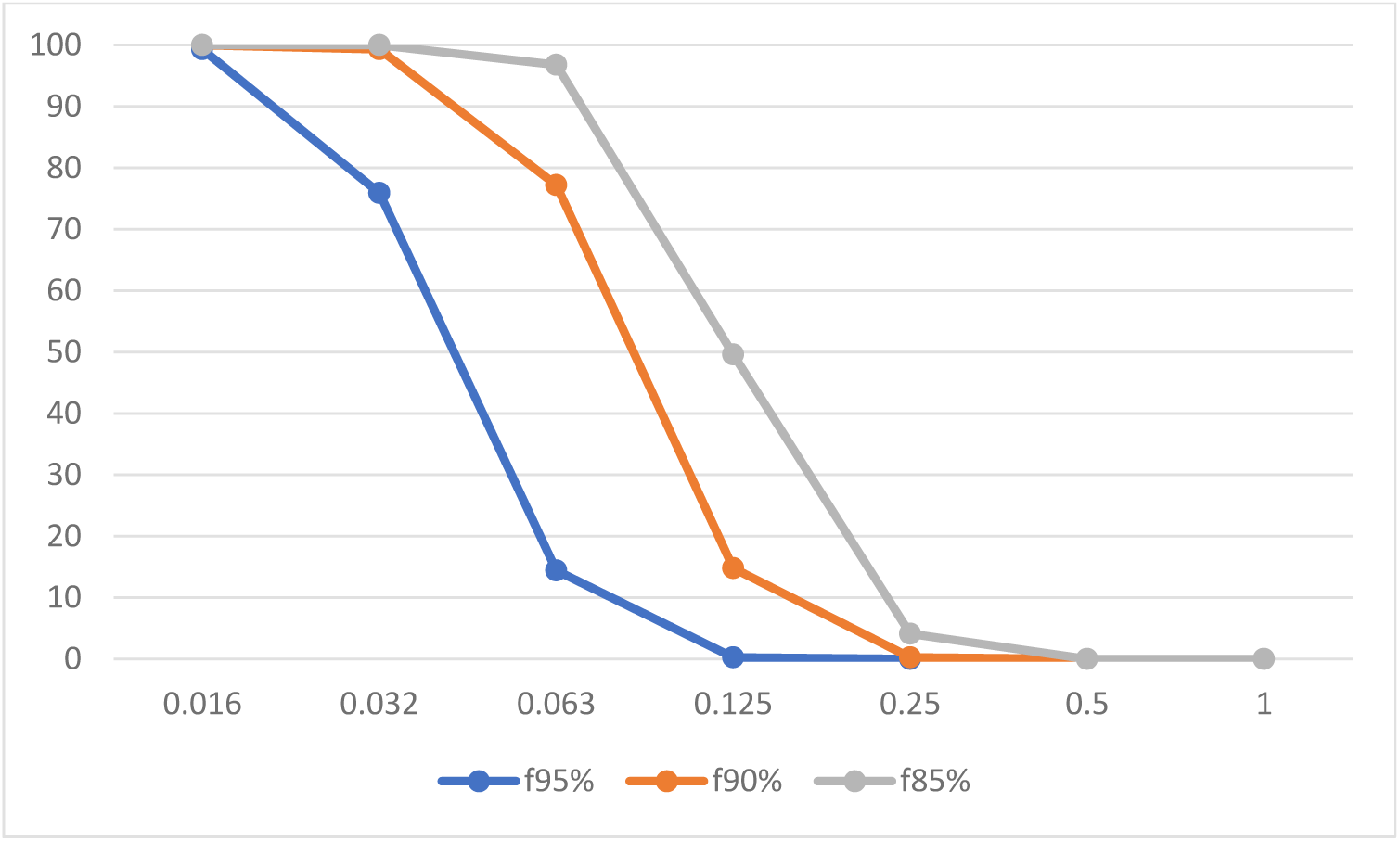

## Appendix 5: Used r packages

library(rxode2)

library(nlmixr2)

library(reshape2)

library(ggplot2)

library(tidyverse)

library(PerformanceAnalytics)

library(psych)

library(dplyr)

library(GGally)

